# Prolonged SARS-CoV-2 Viral Shedding in Patients with COVID-19 was Associated with Delayed Initiation of Arbidol Treatment: a retrospective cohort study

**DOI:** 10.1101/2020.06.09.20076646

**Authors:** Yaya Zhou, Xinliang He, Jianchu Zhang, Yu’e Xue, Mengyuan Liang, Bohan Yang, Wanli Ma, Qiong Zhou, Long Chen, Xiaorong Wang

## Abstract

**Objectives:** Evaluate the risk factors of prolonged SARS-CoV-2 virus shedding and the impact of arbidol treatment on SARS-CoV-2 virus shedding.

**Methods:** Data were retrospective collected from adults hospitalized with COVID-19 in Wuhan Union Hospital. We described the clinical features and SARS-CoV-2 RNA shedding of patients with COVID-19 and evaluated factors associated with prolonged virus shedding by multivariate regression analysis.

**Results:** Among 238 patients, the median age was 55.5 years, 57.1% were female, 92.9% (221/238) used arbidol, 58.4% (139/238) used arbidol combination with interferon. The median time from illness onset to start arbidol was 8 days (IQR, 5-14 days) and the median duration of SARS-CoV-2 virus shedding was 23 days (IQR, 17.8–30 days). SARS-CoV-2 RNA clearance was significantly delayed in patients who received arbidol >7 days after illness onset, compared with those in whom arbidol treatment was started≤7 days after illness onset (HR, 1.738 [95% CI, 1.339–2.257], P < .001). Multivariate regression analysis revealed that prolonged viral shedding was significantly associated with initiation arbidol more than seven days after symptom onset (OR 2.078, 95% CI [1.114-3.876], P .004), more than 7 days from onset of symptoms to first medical visitation (OR 3.321, 95% CI[1.559-7.073], P .002), illness onset before Jan.31, 2020 (OR 3.223, 95% CI[1.450-7.163], P .021). Arbidol combination with interferon was also significantly associated with shorter virus shedding (OR .402, 95% CI[.206-.787], P .008).

**Conclusions:** Early initiation of arbidol and arbidol combination with interferon as well as consulting doctor timely after illness onset were helpful for SARS-CoV-2 clearance.

## Introduction

Outbreak of the novel coronavirus disease (COVID-19) caused by severe acute respiratory syndrome coronaviruse-2 (SARS-CoV-2) in Wuhan City, Hubei Province, China, has been rapidly spreading nationwide and abroad. Up to April 7, 2020, a total of 83,157 COVID-19 cases with 4.03% mortality has been reported in China [1], and there were more than 1,300,000 people with 78,932 deaths have been infected by SARS-CoV-2 globally outside China including more than 180 countries and regions around the world [2]. Building scenarios and strategies including careful management and isolation based on the clinical characteristics and duration of virus shedding of confirmed patients with COVID-19 are important to slow and control transmission of the COVID-19 [3].

Viral replication in respiratory epithelial cells which was associated with virus shedding was the factor contributing to the pathogenesis of SARS-CoV-2 virus infection [4, 5], more importantly, virus shedding was the most important factors in assessing the risk of transmission and guiding decisions regarding isolation of patients with COVID-19. There were many factors contributing to the prolonged respiratory virus other than SARS-CoV-2 virus shedding, such as delayed initiation of antiviral treatment [6-8] and the corticosteroid administration [6, 8], older age [8], major comorbidities [7-9], duration of respiratory symptoms [7] and severity [10]. By now, the therapeutic antivirals for COVID-19 including arbidol, lopinavir–ritonavir, interferon, chloroquine and ribavirin have been recommended [11]. Arbidol, a small indole-derivative molecule, is a broad-spectrum antiviral to treat influenza and other respiratory viral infections [12, 13]. In a recent open-label, randomized, controlled trial study reported that, comparing to standard care, the lopinavir–ritonavir treatment has no more effects on SARS-CoV-2 virus clearance [14]. However, the effects on virus clearance of arbidol for treatment of COVID-19 were unclear.

Here, we conducted a retrospective cohort study of 238 hospitalized patients with laboratory-confirmed COVID-19 in Wuhan Union hospital to identify the risk factors of prolonged SARS-CoV-2 virus shedding, and the effects of arbidol on the SARS-CoV-2 virus clearance as well.

## Methods

### Study design

This is a retrospective observational cohort study of patients with confirmed COVID-19 hospitalized in isolation ward of department of respiratory and critical care medicine, Wuhan Union Hospital from January 15, 2020 to March 15, 2020.

### Data collection

Demographic information, comorbidities, date of symptom onset, symptoms and signs, laboratory test on admission, radiological images and treatment data of patients with COVID-19 were extracted from electronic medical records.

### Laboratory Procedures

All patients were confirmed COVID-19 by real-time reverse transcriptase–polymerase chain reaction assays (RT-PCR) according to WHO interim guidance [15]. To identify SARS-CoV-2 infection, throat swab samples were obtained from all patients at admission and tested using RT-PCR according to the same protocol described previously [16] (Supplementary).

For patients, throat swab specimens were obtained for SARS-CoV-2 PCR re-examination every other day after admission. We defined the duration of viral detection as the number of days from illness onset until the day of the last negative RT-PCR specimen when tested two consecutive negative specimens. In our current analysis, prolonged viral shedding was defined as viral shedding ≥23 days (median duration of SARS-CoV-2 RNA shedding of 238 patients).

### Statistical Analysis

Median values with interquartile range (IQR) were used to describe continuous variables, and absolute or relative frequencies were used to describe categorical variables. We used the Mann Whitney U test for analysis of continuous variables and the χ2 test or Fisher exact test for analysis of discrete variables in bivariate analyses. To identify risk factors associated with prolonged duration of SARS-CoV-2 RNA shedding, we included indicators that were significantly different between the two group except of length of hospital (viral shedding ≥23 vs <23 days) to evaluate the factors associated with virus shedding by univariable regression analysis, and then the significantly contributing factors in univariable regression analysis were included to identify risk factors of prolonged SARS-CoV-2 virus shedding by multivariable logistic regression models. We used Kaplan-Meier survival analysis to estimate the cumulative SARS-CoV-2 RNA–negativity rate and the stratified log-rank statistic to compare the difference of SARS-CoV-2 clearance between patients with or without risk factors.

The statistical analyses and graphics were performed with IBM SPSS 22.0 (SPSS Inc, Armonk, NY) and R 3.6.0 (The R Foundation for Statistical Computing, Vienna, Austria). For all the analyses, P<.05 was considered to be statistically significant and all tests were 2-tailed, unless otherwise indicated.

### Ethical Approval

Ethical approval was waived by the institutional review board of Wuhan Union Hospital since we collected and analyzed all data from the patients according to the policy for public health outbreak investigation of emerging infectious diseases issued by the National Health Commission of the People’s Republic of China.

## Results

### Demographics and Characteristics

There were a total of 242 adult patients with confirmed COVID-19. After excluding 4 non-survivors who tested SARS-CoV-2 RNA positive until death, we included 238 survivors who tested two consecutive SARS-CoV-2 RNA negative, 237 survivors discharged and 1 survivor was still hospitalized before April 7, 2020 in the final analysis. The median age was 55.5 years (IQR, 35-67.3 years), and 136 patients (57.1%) were female. The most common symptoms on admission were fever and cough, followed by fatigue. 88 patients (37.0%) had comorbidities. 92.9% (221/238) used arbidol, 61.7% (147/238) used interferon. Among them, arbidol monotherapy in 82 (34.4%); arbidol combination with interferon in 139 (58.4%); interferon monotherapy in 3 (1.3%); interferon and lopinavir/ritonavir in 5 (2.1%); and lopinavir/ritonavir monotherapy in 1 (0.4%); and miss in 8 (3.4%) (Table 1).

**Table 1.**
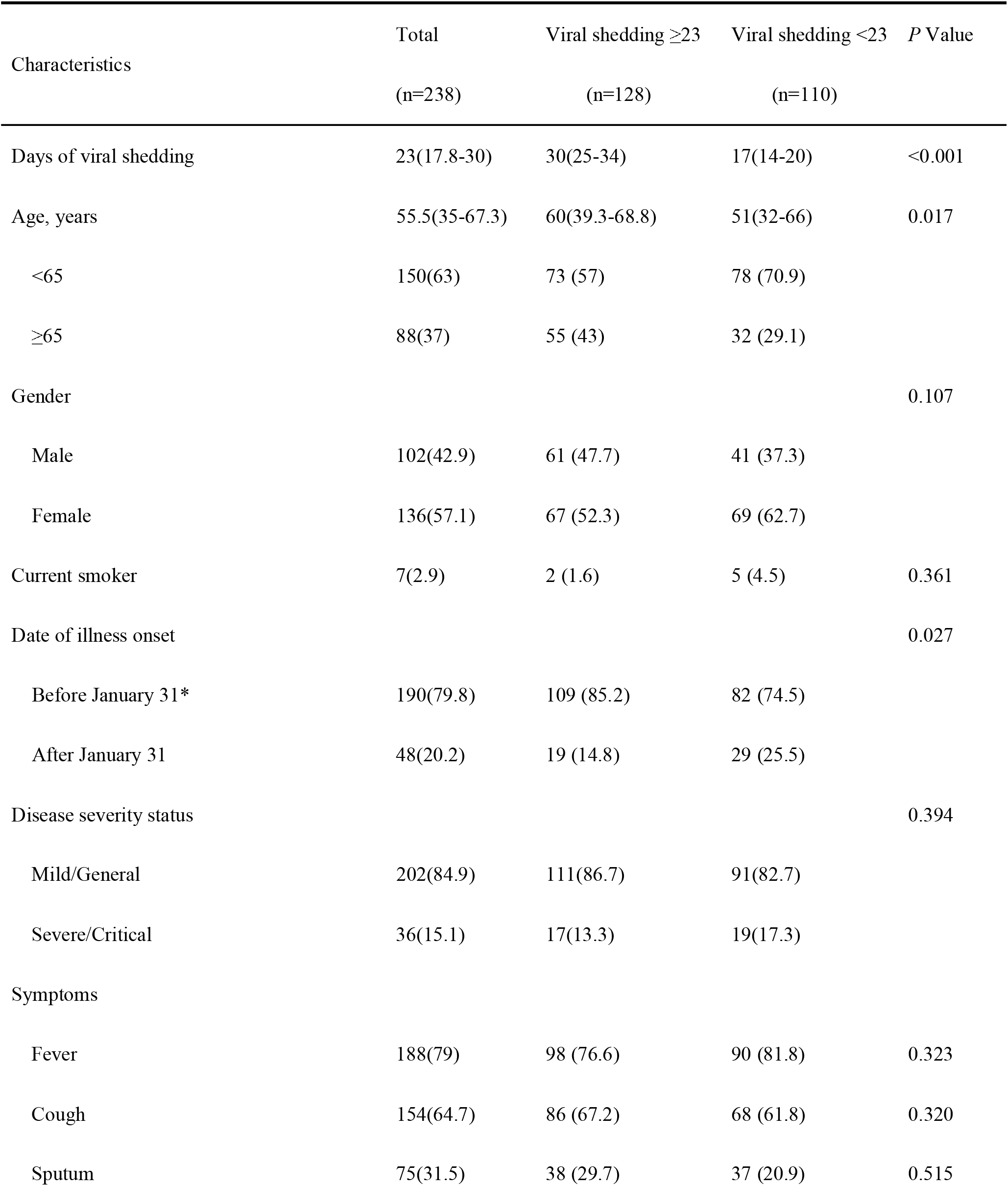
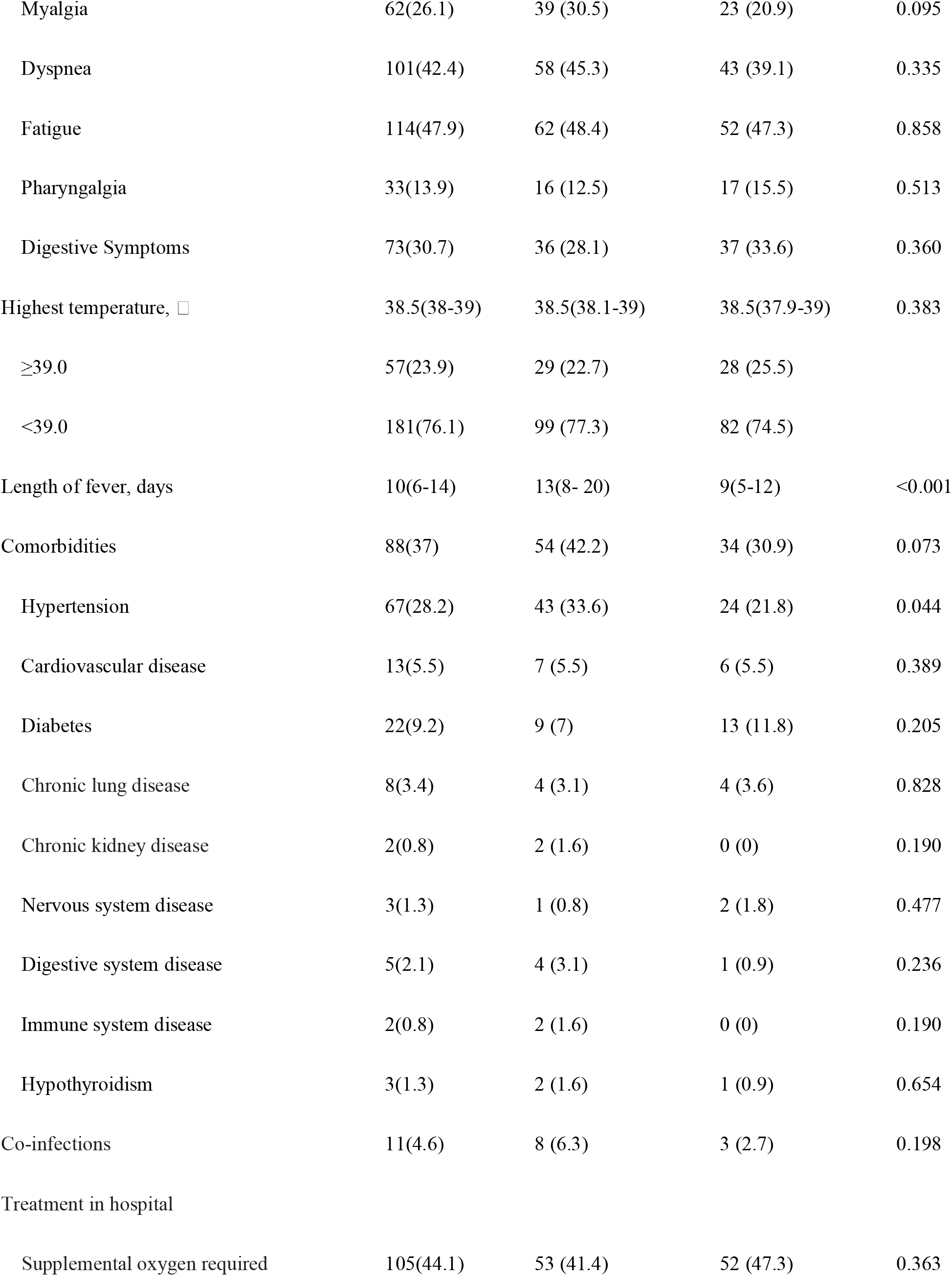
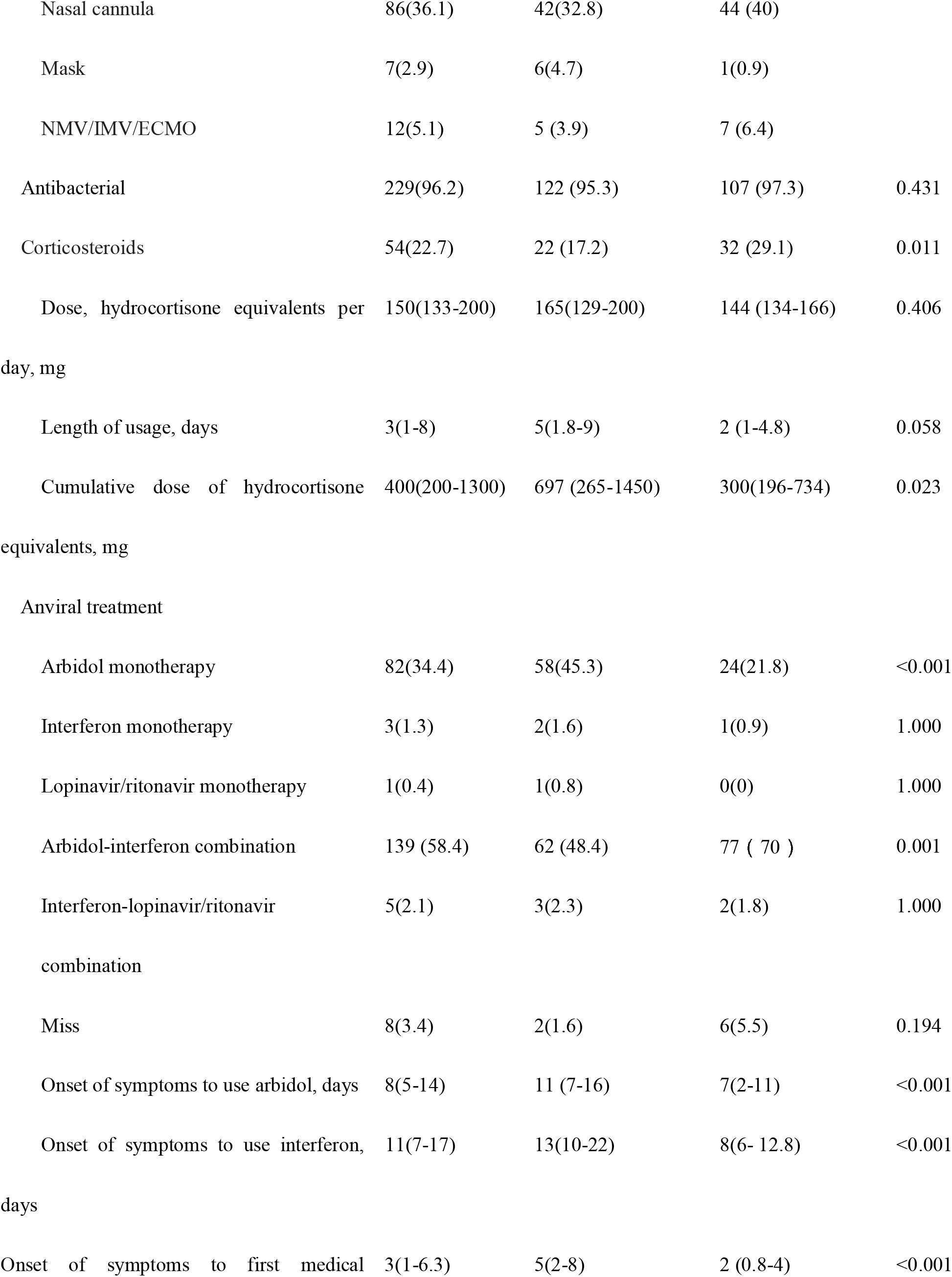
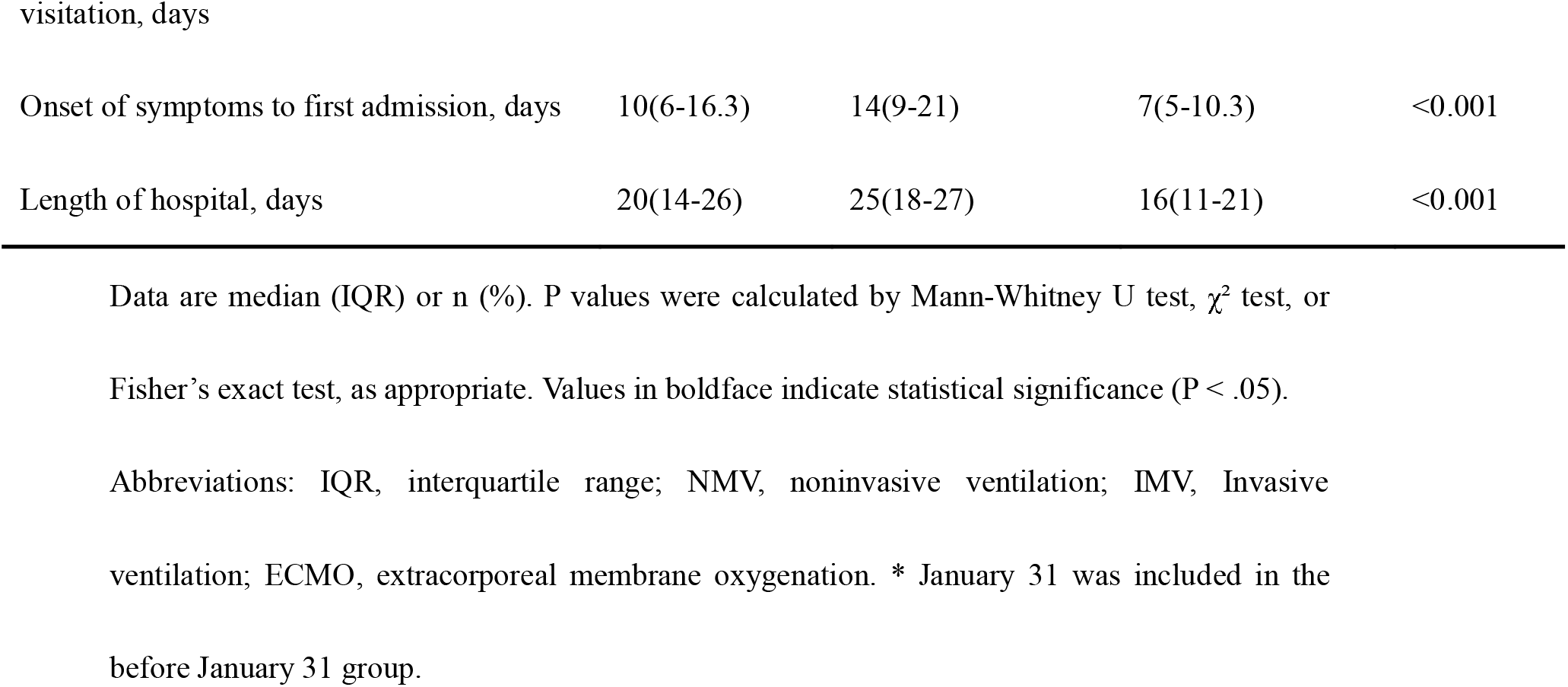
Demographic and Clinical Characteristics of Patients With COVID-19 on Admission

### SARS-CoV-2 Viral Shedding and Comparison of Characteristics of Patients with Prolonged and Short Viral Shedding

The median duration of viral shedding was 23 days (IQR, 17.8–30 days) from illness onset. The shortest observed duration of viral shedding was 9 days, whereas the longest was 51 days (Figure 1).

**Figure 1.**
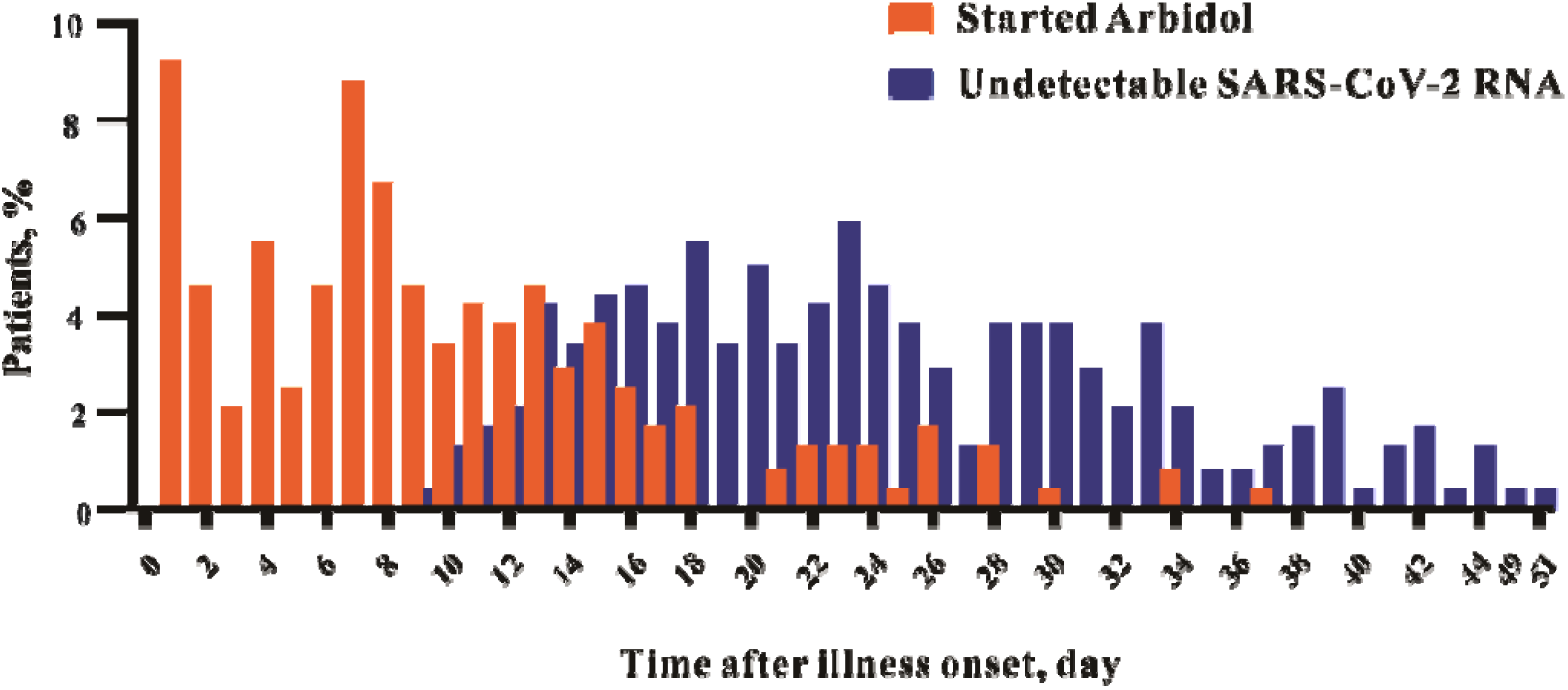
Distribution of arbidol treatment and proportion of patients with undetectable SARS-CoV-2 RNA by day after onset of symptom. Among 238 patients, 221 patients (92.9%) received arbidol treatment, 17 patients (7.1%) did not receive arbidol treatment.

The data of patients with viral shedding <23 days and ≥23 days groups were compared (Table 1). Compared with patients with viral shedding <23 days, patients with prolonged viral shedding were older (p=.017), a higher proportion of hypertension (33.6%, p=.044), longer time of fever (p<.001), longer time of hospital (p<.001) and later of first medical visitation (p<.001). More patients with prolonged viral shedding had onset symptoms before January 31 compared with those with viral shedding <23 days (85.2%, p=.027). Less patients with prolonged viral shedding received corticosteroids treatment (17.2%, p=.011), but the cumulative doses of corticosteroid usage were significantly higher (p=.023) than that of patients with virus shedding less than 23 days. As summarized in Supplementary Table 1, the time from illness onset to arbidol or interferon treatment, elevated C-reactive protein (CRP), elevated interleukin-4 (IL-4) and interleukin-6 (IL-6), elevated D-dimer and more lobes lesion in lung CT images were significantly associated with the prolonged viral shedding of COVID-19. Compered to prolonged viral shedding group, more patients received arbidol combination with interferon treatment (p=.001) in patients with virus shedding <23 days (Table 1).

### Risk Factors for Prolonged Viral Shedding

The results of univariable regression analysis demonstrated that age older than 65 years, illness onset before January 31, the time from illness onset to first medical visitation, hypertension, arbidol combination with interferon, lobe lesions in lung computed tomography (CT) images, and the time from illness onset to arbidol or interferon initiation were significantly associated with the duration of SARS-CoV-2 RNA shedding by univariable regression analysis. Other indicators including cumulative dose of corticosteroids was not associated with prolonged viral shedding significantly (Table 2).

**Table 2.** Univariate and Multivariate Logistic Regression Analysis for Prolonged Viral Shedding of Patients with COVID-19

|  | Univariate |  | Multivariate |  |
| --- | --- | --- | --- | --- |
|  | OR (95% CI) | P-value | OR (95% CI) | P-value |
| Age≥65 years old | 1.836 (1.070-3.152) | 0.027 | 1.075 (0.534-2.166) | 0.839 |
| Male sex | 1.532 (0.912-2.575) | 0.107 |  |  |
| Date of illness onset before Jan. 31 | 2.054 (1.077-3.919) | 0.029 | 3.223 (1.450-7.163) | 0.004 |
| Onset of symptoms to first medical<br>visitation >7 days | 5.588(2.787-11.202) | <0.001 | 3.321 (1.559-7.073) | 0.002 |
| Hypertension | 1.813 (1.012-3.246) | 0.045 | 1.332 (0.656-2.707) | 0.428 |
| Cumulative dose of corticosteroids | 1.000(1.000-1.001) | 0.784 |  |  |
| Starting arbidol >7 days from illness onset | 3.062(1.791-5.234) | <0.001 | 2.078 (1.114-3.876) | 0.021 |
| Interferon and arbidol combination | 0.403(0.236-0.688) | 0.001 | 0.402 (0.206-0.787) | 0.008 |
| Starting interferon >7 days from illness<br>onset | 4.643(2.004-10.755) | <0.001 | 1.766 (0.685-4.554) | 0.239 |
| CRP >8.0mg/L | 0.803(0.474-1.359) | 0.413 |  |  |
| IL-4 >3.2pg/ml | 3.353(0.911-12.344) | 0.069 |  |  |
| IL-6 >2.9pg/ml | 1.382(0.450-4.242) | 0.572 |  |  |
| D-dimer >0.5μg/L | 1.577(0.943-2.636) | 0.082 | | |
| More lung lobes lesion in chest CT images | 2.448(1.366-4.387) | 0.003 | 1.726 (0.871-3.419) | 0.118 |

Abbreviations: CI, confidence interval; OR, odds ratio; CRP, C-reactive protein; IL-4, interleukin-4; IL-6, interleukin-6; CT, computed tomography.

The multivariable logistic regression showed the time from illness onset to arbidol initiation more than 7 days, first medical visitation after illness onset more than 7 days and illness onset before January 31 were independent factors associated with the duration of SARS-CoV-2 RNA shedding. Importantly, arbidol combination with interferon was also significantly associated with shorter virus shedding (Table 2).

### Impact of Delayed Arbidol Treatment and Arbidol Combination Regimens on Virus Shedding

The median time from illness onset to arbidol treatment initiation was 8 days (IQR, 5-14 days). and the median duration of SARS-CoV-2 virus shedding was 20 days (IQR, 16-25 days) in the patients with initiation of arbidol ≤7 days and 28 days (IQR, 21-33) in patients with initiation arbidol >7 days. SARS-CoV-2 RNA clearance was significantly delayed in patients who received arbidol treatment beginning >7 days after illness onset, compared with those in whom arbidol treatment was started≤7 days after illness onset (HR, 1.738 [95% CI, 1.339-2.257], P <.001, Figure 2A). The characteristics of patients of the two groups were summarized in Table 3. Age was significantly different in two groups, so we further compared characteristics with patients who were less than 65 years in two groups, the results were showed in Supplementary Table 2.

**Figure 2.**
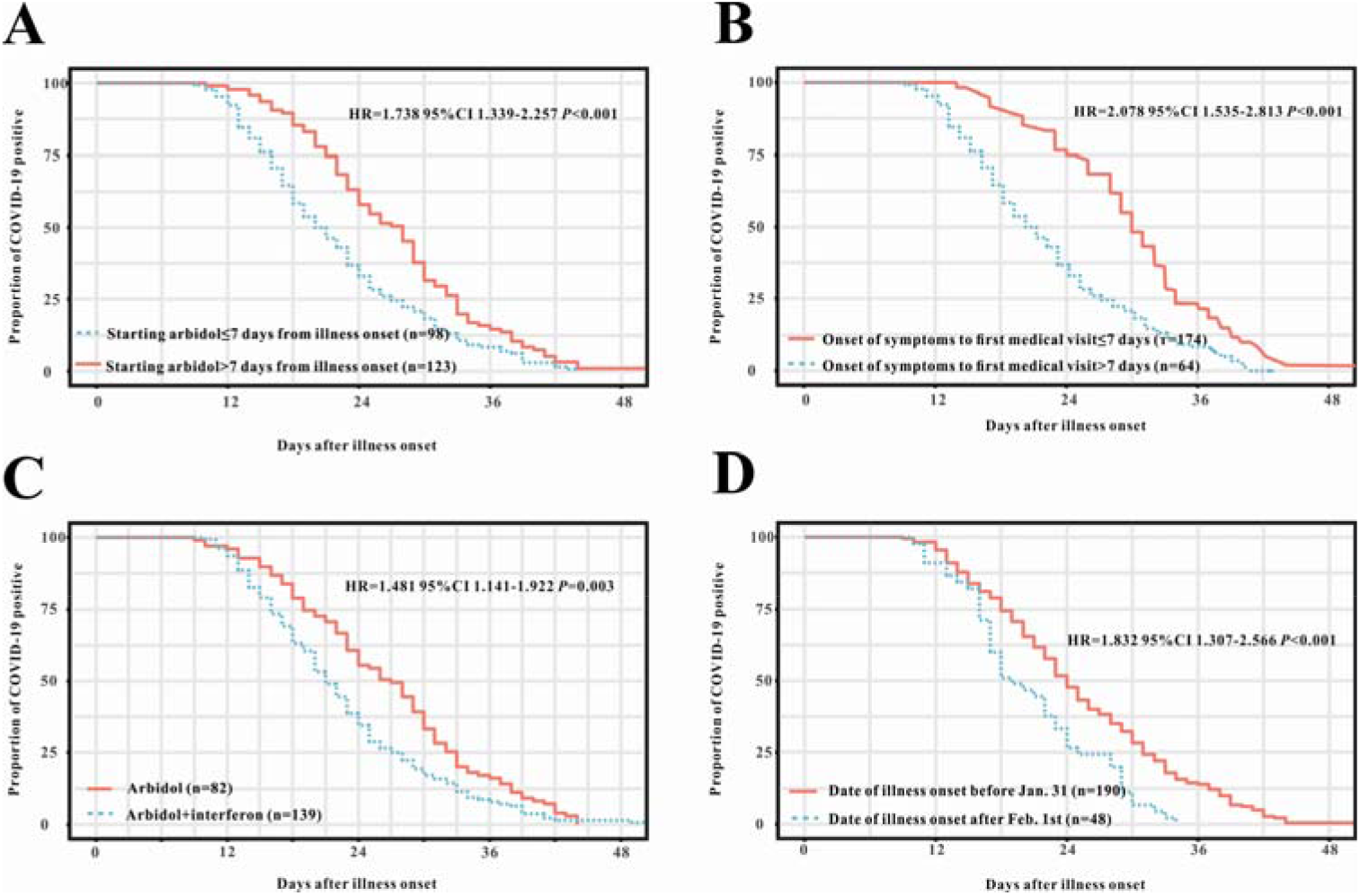
A. Cumulative proportion of between patients who started arbidol therapy ≤7 days and >7 days after illness onset with detectable SARS-CoV-2 RNA, by day after onset of illness. B. Cumulative proportion of patients who firstly visited medical staff ≤7 days from symptoms onset versus >7 days who had detectable SARS-CoV-2 RNA, by day after onset of illness. C. Cumulative proportion of patients treated with arbidol versus combination of arbidol and interferon who had detectable SARS-CoV-2 RNA, by day after onset of illness. D. Cumulative proportion of patients who onset illness before Jan. 31 versus after Feb. 1 who had detectable SARS-CoV-2 RNA, by day after onset of illness. HR, hazard ratio; CI, confidence interval. Jan. 31 was included in before January 31 group and Feb. 1 was included in after Feb. 1 group.

**Table 3.**
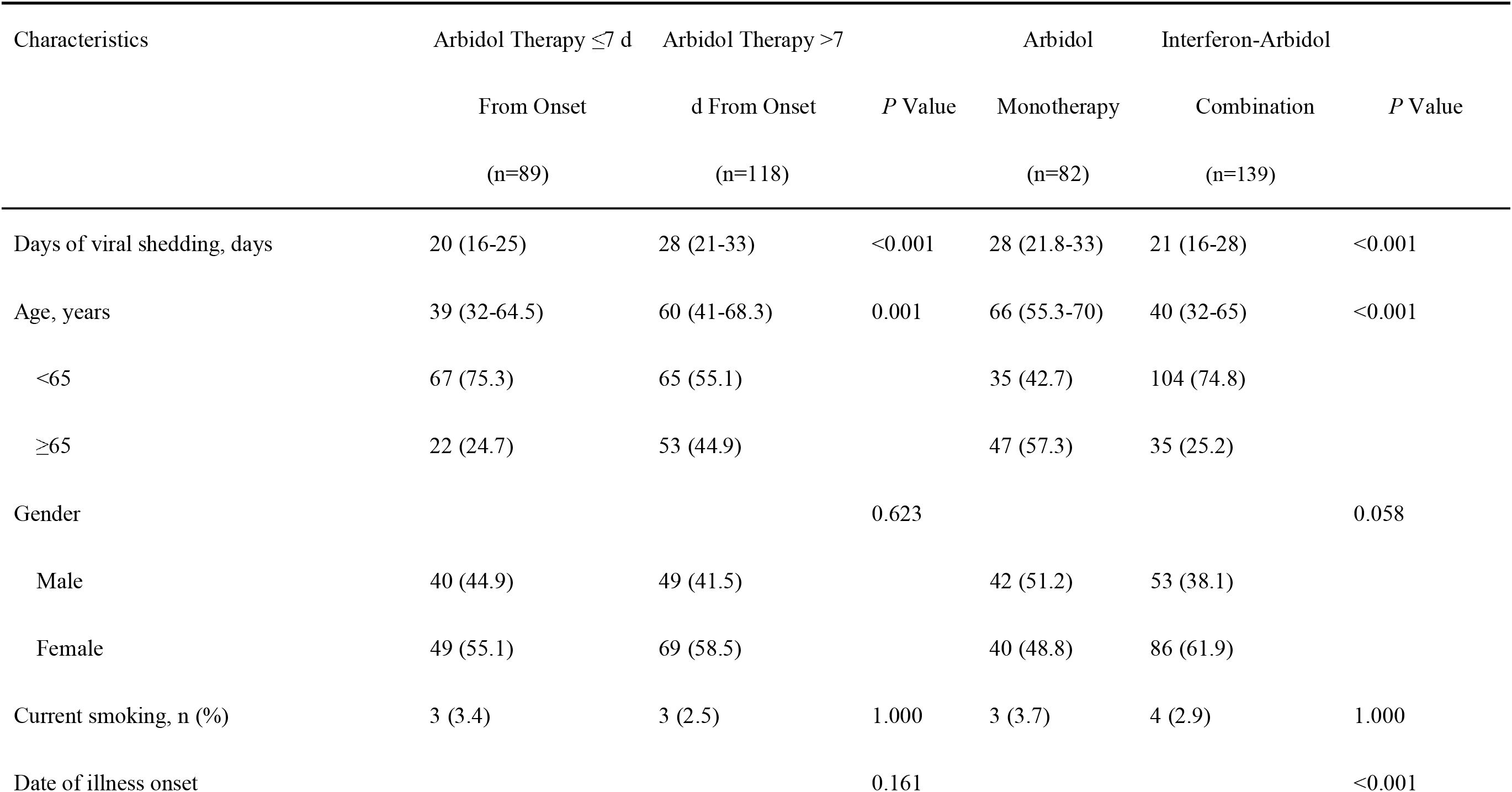
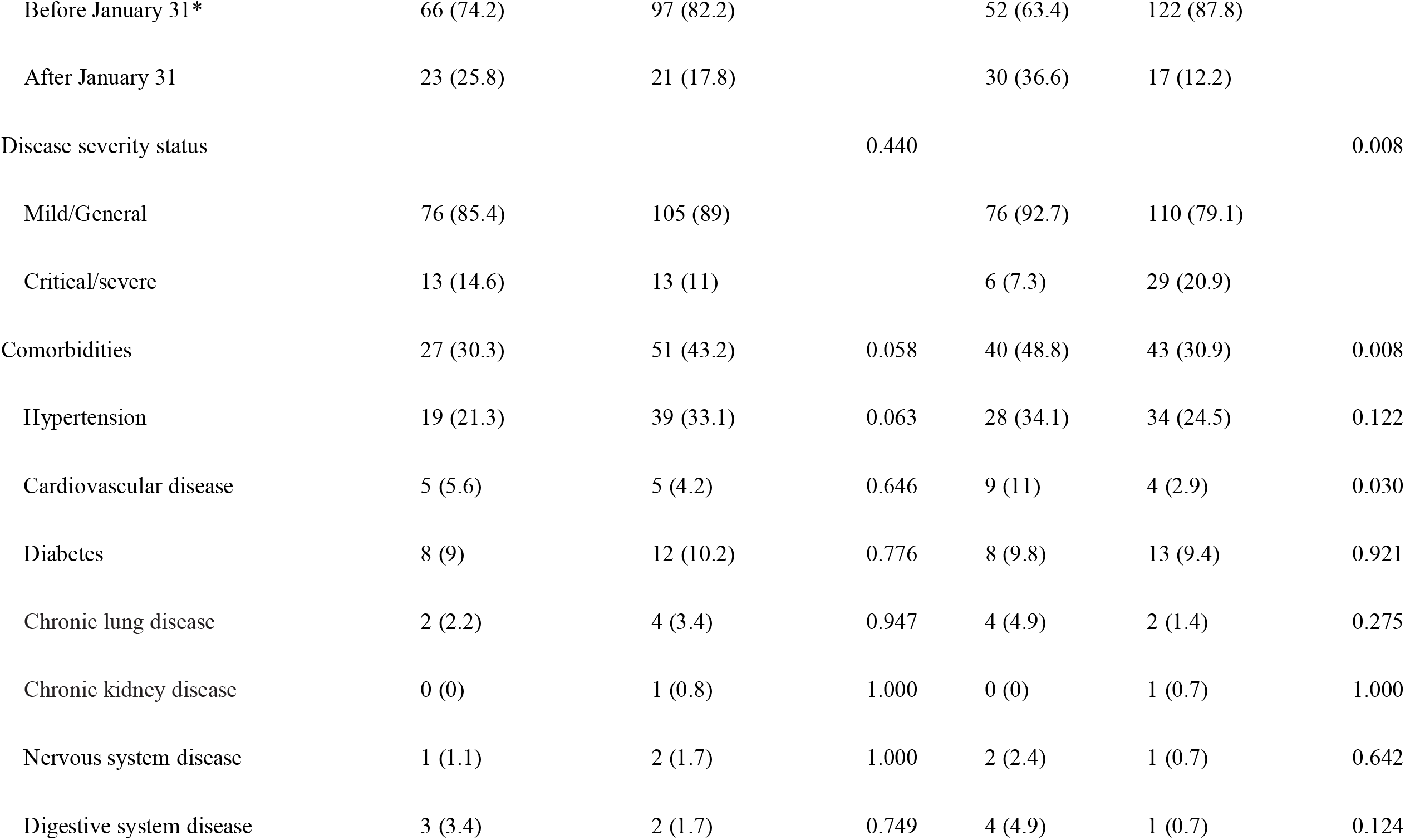
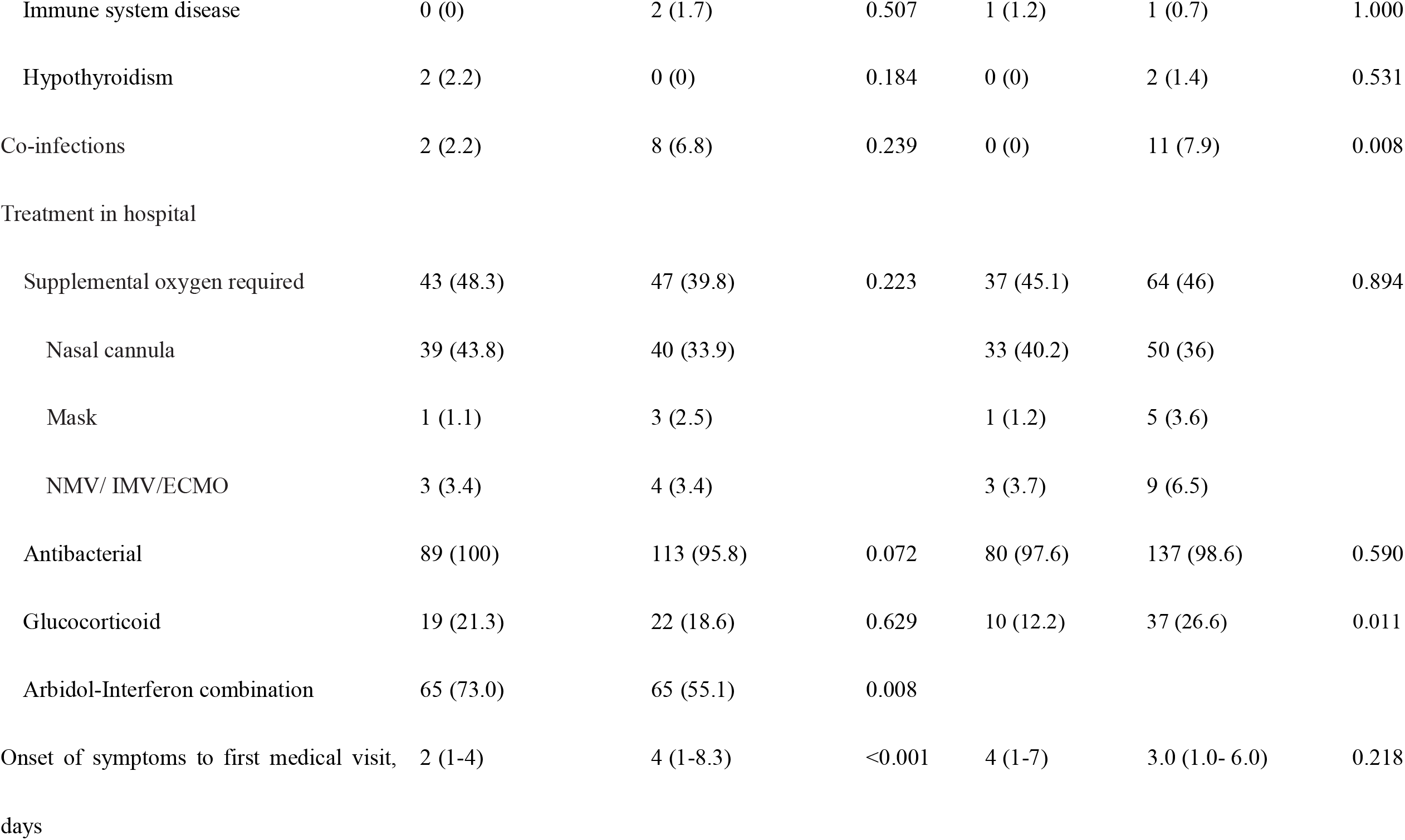
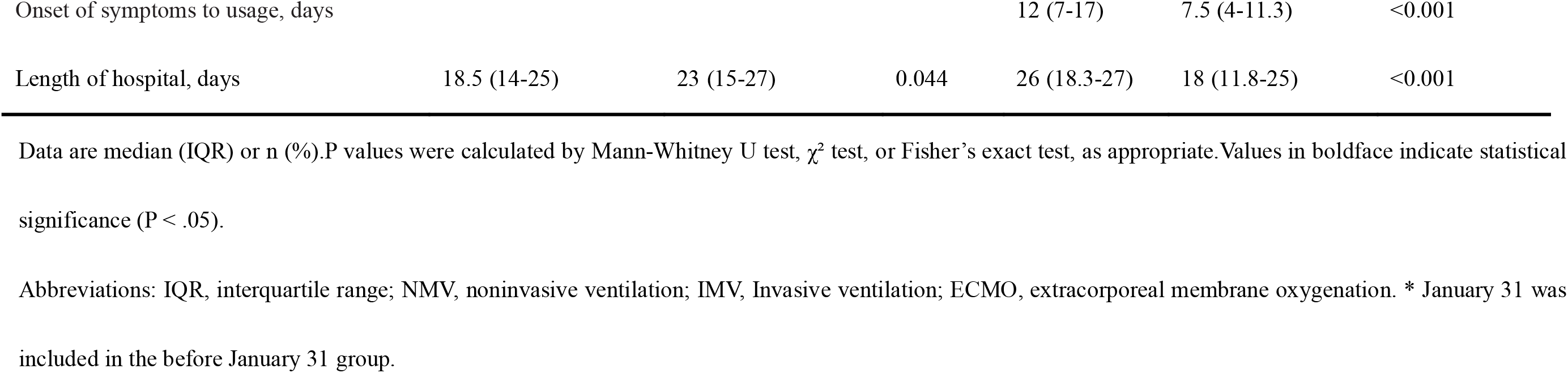
Comparation of Demographic and Clinical Characteristics between Patients With COVID-19 Who Treated With Arbidol

Comparing to arbidol monotherapy, the median durations of SARS-CoV-2 shedding of patients with arbidol-interferon combination treatment (21 days, IQR, 21.8-33 days) was significantly shorter than that of arbidol monotherapy (28 days, IQR, 16-28 days) (P <.001). The delayed virus clearance was also seen in patients in arbidol monotherapy, in compared with those who received arbidol-interferon combination therapy in Kaplan-Meier survival analysis (Figure 2C). The main characteristics of patients who received arbidol monotherapy and arbidol-interferon combination are summarized in Table 3.

## Discussion

In the current cohort study of 238 hospitalized patients with COVID-19, we found that the duration of SARS-CoV-2 RNA shedding was very long and initiation of arbidol within seven days after illness onset as well as combination with arbidol and interferon were helpful for SARS-CoV-2 RNA clearance. We also found that the time from onset of symptoms to first medical visitation more than 7 days as and illness onset before Jan.31, 2020 as independent risk factors for prolonged SARS-CoV-2 RNA detection.

We detected SARS-CoV-2 RNA in the respiratory tract for a median duration of 23 days with a longest one was 51 days in an old man with severe COVID-19, which was a little longer than 20 days reported by an early study [17]. The much longer virus shedding may correlate with the SARS-CoV-2 pandemic around the world, and demonstrates more time may be required to quarantine the patients with COVID-19 and the length of antiviral treatment may be longer. The prolonged shedding duration of highly pathogenic avian influenza A (H5N1) and A (H1N1) pdm09 virus shedding were correlated with disease severity [6, 10, 18, 19]. Compared to patients with virus shedding less than 23 days, more patients with prolonged virus shedding has abnormally higher valued of D-dimer, IL-4, IL-6, CRP, more lobes lesion in lung images, older age and more patients with hypertension. Previous studies reported that the D-dimer and older age were risk factors of death and acute respiratory distress syndrome (ARDS) in patients with COVID-19 [17, 20]. Additionally, higher inflammation markers, more lobes lesion in lung images and comorbidities associated with severity of COVID-19 [11]. Those results combined with our present findings demonstrated that patients with prolonged virus shedding are more likely to develop to severe stations.

Antiviral therapy especially early initiated antiviral therapy has been proven to reduce the viral shedding period of seasonal and epidemic influenza as well as H7N9 avian influenza [6, 7, 21-23]. In this retrospective study, most patients (221/238, 92.9%) used arbidol, and the findings demonstrated that initiation of arbidol within 7 days after illness onset was associated with a shorter duration of SARS-CoV-2 RNA shedding. Multivariate regression analysis revealed delayed arbidol antiviral therapy was an independent risk factor of the prolonged SARS-CoV-2 RNA shedding. The antiviral mechanism of arbidol involves inhibition of virus-mediated fusion with target membrane and a resulting block of virus entry into target cells was revealed to effectively inhibit SARS-CoV in vitro [24]. Arbidol showed tendency to improve discharging rate and reduce mortality of patients with COVID-19 [25]. These results combined with our present findings suggest that arbidol has a therapeutic effect on SARS-CoV-2 infections, however, further research is needed to clarify the effects of additional arbidol treatment for COVID-19.

Interferon, which was reported to inhibit SARS-CoV reproduction in vitro [26], is also a broad-spectrum antiviral and is recommended to treat coronavirus infection combining with other antivirals [27]. In the present study, about 147 (61.8%) of patients used interferon, among them, up to 94.6% (139/147) was used in combination with arbidol. Our findings showed arbidol combination with interferon can help to clear SARS-CoV-2 RNA. The effects of arbidol combination with interferon treatment for COVID-19 worth further study.

The usage of corticosteroids in patients with COVID-19 was controversial [28]. Most observational studies have reported that use of corticosteroids was associated with persistent viral shedding [6, 7, 29]. In the present study, 22.7% used corticosteroids and most of them used in a short duration, and it was not associated with prolonged SARS-CoV-2 RNA shedding, that are in accordance with the findings in a retrospective study in china [30]. However, the length and cumulative dose of corticosteroids usage were higher in prolonged shedding group, which demonstrated the long-term use of corticosteroids to a certain dose has a tendency to delay viral clearance.

Our present finding showed that the time from onset of symptoms to first medical visitation more than 7 days as an independent risk factor for prolonged SARS-CoV-2 RNA detection, suggesting that consulting a doctor as soon as possible after illness onset helps to clear SARS-CoV-2 (Table 2, Figure 2B). Interestingly, illness onset before Jan.31 was also associated with significant differences in SARS-CoV-2 RNA shedding, compared with illness onset after Jan.31 (Table 2, Figure 2D). This may be associated with shortage of medical resources due to the outbreak of COVID-19 in Wuhan and a large number of patients could not see a doctor and received effective treatment timely in January, 2020. Additionally, the genomic mutation during the virus transmissibility may correlate with the SARS-CoV-2 virus shedding.

Some limitations of this study should also be acknowledged. First, the retrospective single-center design leads to missing data and unavoidable biases and the sample size was relatively small. Second, owing to the small number of patients who initiated arbidol treatment 2–3 days of illness onset, we were not able to assess the effect of earlier initiation of arbidol treatment. Despite these limitations, the study was designed to reflect the ‘real life’ clinical situation. Clinical information was meticulously gathered using standard protocols by admitted medical team. This has important implications for both patient isolation decision making and guidance around the length of antiviral treatment and the choice of antivirals.

To the best of our knowledge, this is the first retrospective cohort study among patients with COVID-19 to assess the risk factors of prolonged virus shedding and the effects of arbidol on SARS-CoV-2 shedding. We found that initiation of arbidol within seven days after illness onset as well as arbidol combination with and interferon were helpful for clearance of SARS-CoV-2 RNA. Additionally, prolonged SARS-CoV-2 RNA shedding was independently associated with delayed medical visitation and correlated with the illness onset day (before or after 31, Jan). These results reinforce consulting a doctor and receiving treatments as soon as possible after illness onset helps to SARS-CoV-2 clearance in patients with COVID-19.

## Data Availability

I state that the all data referred to in the manuscript and note links below is available.

## Acknowledgements

We acknowledge all health-care workers involved in the diagnosis and treatment of patients in Wuhan.

## Authors Contributors

XW had the idea for and designed the study, XW had full access to all data in the study and take responsibility for the integrity of the data and the accuracy of the data analysis. XW, YZ contributed to the drafting of the manuscript and data analysis. All authors contributed to data acquisition, data analysis, or data interpretation. The final version had been reviewed and approved by all authors.

